# Systematic review of genome wide association studies (GWAS) of epilepsy identifies common risk variants and associated genes

**DOI:** 10.1101/2023.08.31.23294726

**Authors:** S. Jacobs, O. Wootton, V Ives-Deliperi, L.M Tucker, D.J Stein, S. Dalvie

**Affiliations:** Neuroscience Institute, Department of Mental Health and Psychiatry, University of Cape Town, South Africa; Constantiaberg Medi Clinic, Cape Town, South Africa; Neuroscience Institute, Division of Neurology, University of Cape Town, South Africa; Division of Human Genetics, Department of Pathology, University of Cape Town, South Africa

**Keywords:** Single nucleotide polymorphism (SNP), sodium channels, genome wide association studies, *BRAP*, *ALDH2*

## Abstract

**Background:** Epilepsy is one of the most prevalent neurological disorders, affecting 50 million individuals worldwide. The aetiology of epilepsy is known to have genetic contributions, yet headline results obtained from genome-wide association studies (GWAS) of epilepsy have not always been consistent. We undertook a systematic review of all the findings from this work in order to identify risk variants for epilepsy.

**Methods:** This systematic review was conducted in accordance with the updated PRISMA protocol. EbscoHost, PubMed, Scopus, Web of Science and Primo were searched between the years 2010 – 2024. The quality of each of the studies was evaluated using the Q-Genie tool.

**Results:** 11 studies were included for full extraction, analysis, and quality assessment. Across the identified studies, 79 SNPs, located in 64 genes, were significantly associated with epilepsy at the genome-wide level. The majority of the variants were intronic and intergenic, with the well documented *SCN1A* the most widely reported gene involved across studies. Two SNPs, rs2292096 and rs149212747, linked respectively to focal epilepsy (FE) and status epilepticus, were exclusively identified in individuals of Asian ancestry, alongside an Asian-exclusive synonymous variant (rs3782886) in *BRAP* and a missense variant (rs671) in *ALDH2*.

**Conclusions:** Genes which encode for ion and transport channels, transcription factors, ubiquitin ligase and transporter proteins were identified as potentially involved in the aetiology of epilepsy disorders. The review identified one missense and one synonymous variant which deserve further exploration. Future research should also include populations of more diverse ancestries as this may reveal unique and important epilepsy-associated genes.

## 1. Introduction

Epilepsy is a neurological disorder, characterized by a combination of clinical and electroencephalographic (EEG) features and affects approximately 50 million individuals worldwide ^1^. Epilepsy has four main classification types, namely, i) focal onset, ii) generalized onset, iii) combined focal and generalized onset iv) unknown epilepsy onset and v) epilepsy syndromes ^1,2,3^. Approximately 80% of epilepsy cases occur in low and middle-income countries (LMICs), with a higher incidence than in high income countries (HICs) ^4^. Specifically, the incidence of epilepsy in HICs is 45 per 100 individuals per year, in comparison to 81.7 per 100 per year in LMICs ^5^.

In 2017, the International League Against Epilepsy (ILAE) taskforce updated the classification system for epilepsy to include etiology, age, comorbidities, and genetic associations ^1^. Collectively, molecular genetics, neuroimaging and animal model studies have shown the impact of genetic variations within the human brain, supporting the idea that specific mutations may influence long-term brain maturation during neurodevelopment, resulting in changes to both the structure and function of the brain. These alterations could contribute to an increased susceptibility to seizures and, ultimately, the development of epilepsy and associated comorbidities ^6^.

Many forms of epilepsy have a genetic component, with heritability differing per epilepsy type ^7^. Developmental and epileptic encephalopathy (DEE) is the most well researched epilepsy type reported to be due to genetic etiology, with over 100 reported genes including *SCN1A, SCN2A, KCNQ2* and *STXBP* ^8^. With the 2017 updated classification by the ILAE, the taskforce advised that the term genetic generalized epilepsy (GGE) may be used to classify and group a broader range of epilepsy syndromes which are characterized by clinically generalized seizure types associated with generalized, epileptiform spike-wave discharges, which include generalized epilepsy (GE) syndromes. GE has an estimated heritability of 31%^9^. Idiopathic generalized epilepsy (IGE), is a sub-type of GGE and includes childhood absence epilepsy (CAE), juvenile absence epilepsy (JAE), juvenile myoclonic epilepsy (JME), and generalized tonic-clonic seizures alone (GTCA) ^1,9^.

Over 95% of epilepsy patients are diagnosed with common forms of epilepsy, which include IGE, temporal and frontal lobe epilepsy (TLE/FLE) and benign epilepsy with centrotemporal spikes (BECTS) ^9^. These forms of epilepsy may involve multiple genetic variants, interacting with environmental factors ^10^. Such genetic variants have been identified by genome-wide association studies (GWASs) ^11^. Indeed, over the last decade, multiple GWAS for epilepsy have been conducted, mainly in European-ancestry and Asian-ancestry individuals ^12^. However, headline findings from different studies have not always been consistent. Here we undertook a systematic review of findings from GWASs of epilepsy in order to identify genetic risk variants for this condition.

## 2. Materials and Methods

This systematic review was registered on PROSPERO in March 2022 (ID: CRD42022308427). The PRISMA ^13^ workflow (**Figure 1**) was used to identify all relevant studies published between the years 2010-2024. We had selected the time period based on the fact that the first GWAS on epilepsy was published in 2010. Filters were included on all searches, where applicable, to identify results for studies that included only human participants and were published in English. Duplicates were removed using Endnote’s native tool before exporting into Covidence for title, abstract and full text screening ^14^. Studies were included if they were published in English, were primary GWAS, used human participants and were published between 2010-2022. Studies were excluded if they assessed variants other than single nucleotide polymorphisms (SNPs). We included GWAS performed for any epilepsy type and did not limit our search to specific sub-types. EbscoHost, PubMed, Scopus, Web of Science and the University of Cape Town’s (UCT) Primo were selected as the relevant databases for searching. Search strings combining all possible MeSH terms, as well as search strings combining only two terms at a time were run. Search strings included variations of “(GWAS)”, “((genome wide association stud*)/(analys*))”, “(genome wide stud*)/(analys*))”, “(epileps*)” and “(seizure disorders)”. An updated search was done across all mentioned databases in February 2024, after a recent GWAS publication by the International League Against Epilepsy in August 2023.

**Figure 1.**
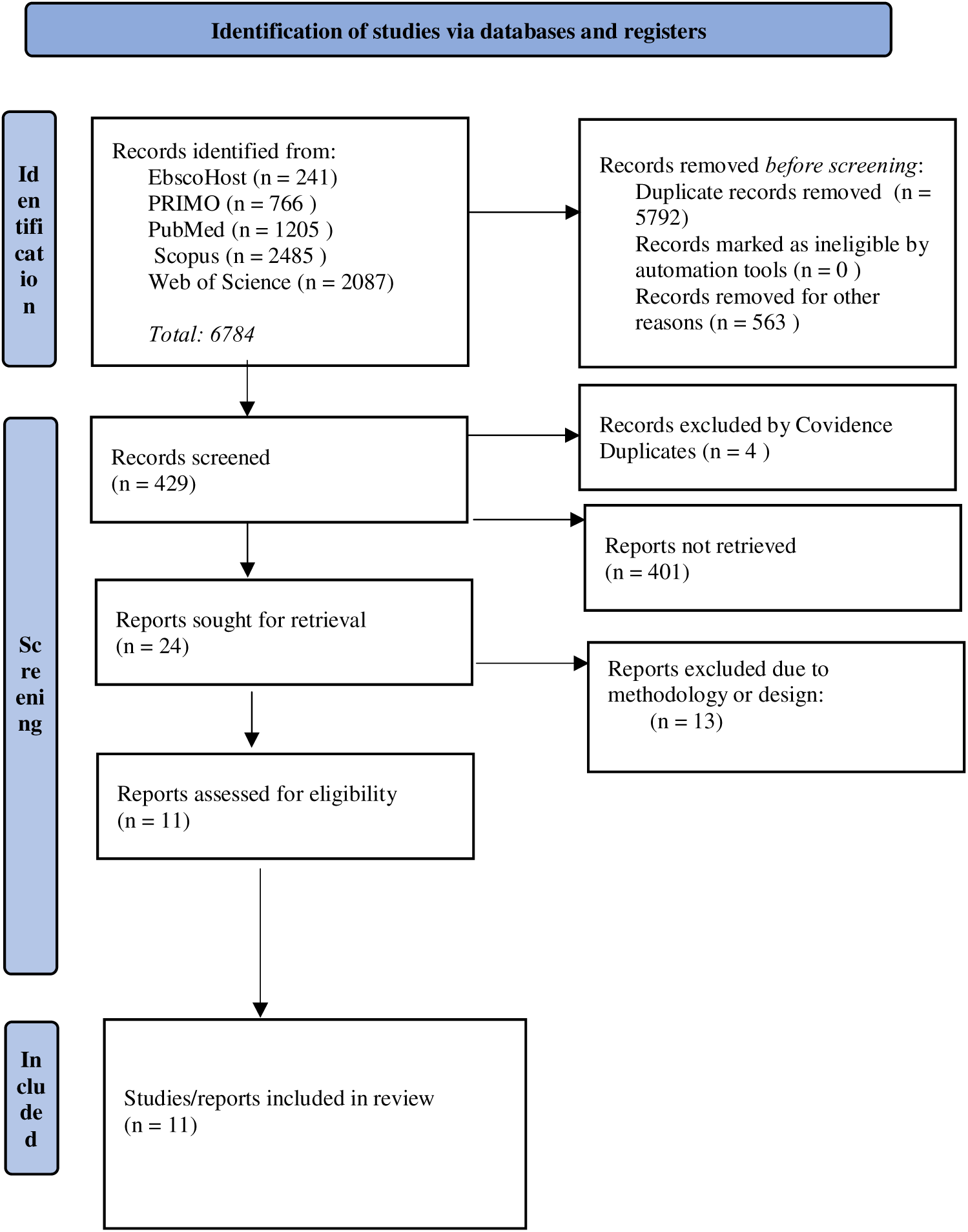
Overview of screening process.

Study characteristics, which included participant number, number of significant SNPs, sequencing platform, epilepsy type, method of diagnosis and ancestry of participants were tabulated (**supplementary materials, Table S1.1**). All SNPs reaching genome-wide significance (p < 5 x 10^-8^), along with their minor allele frequencies (MAF), effect allele, quality score and associated epilepsy type, were extracted (**Table 1**).

**Table 1.**
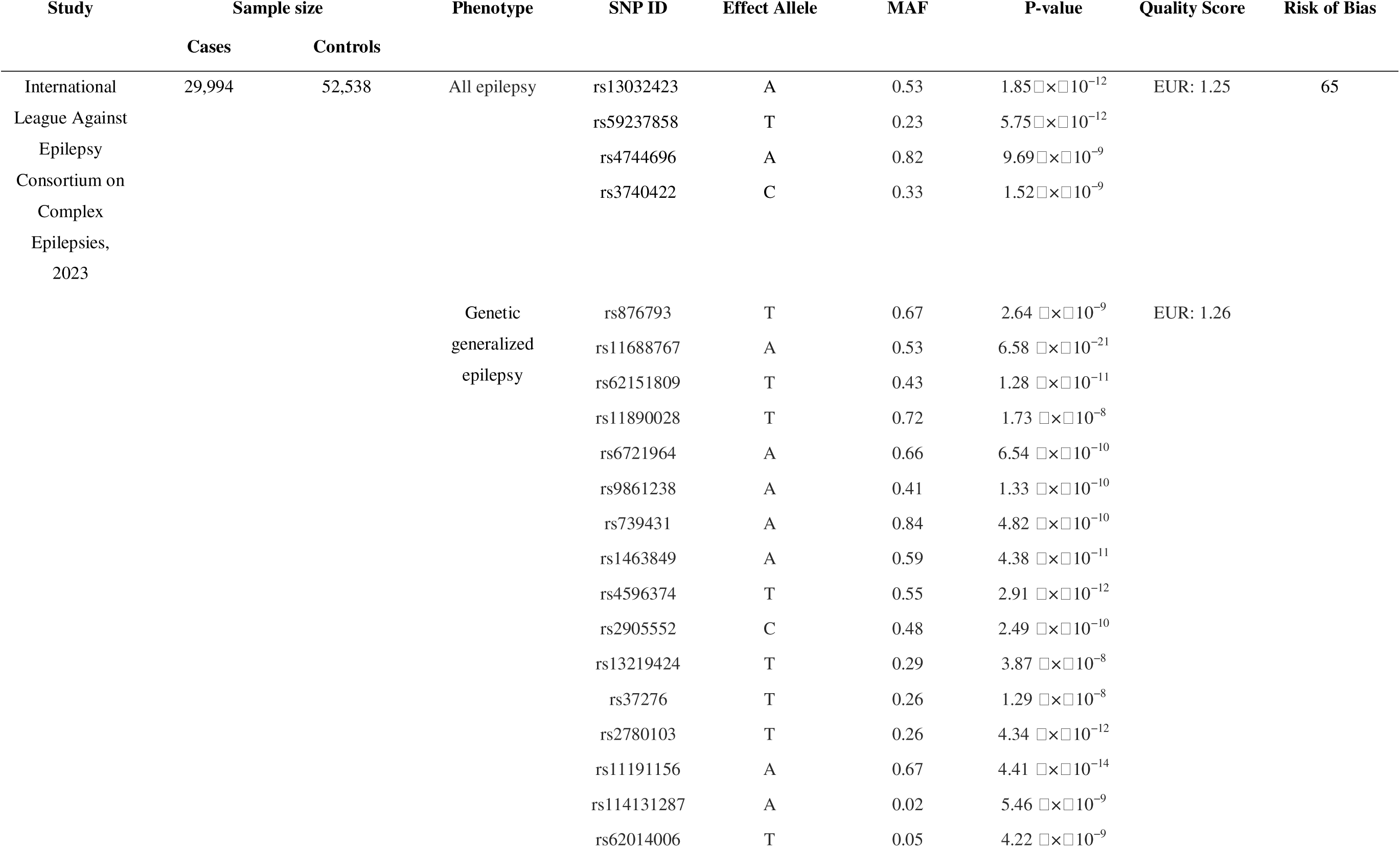
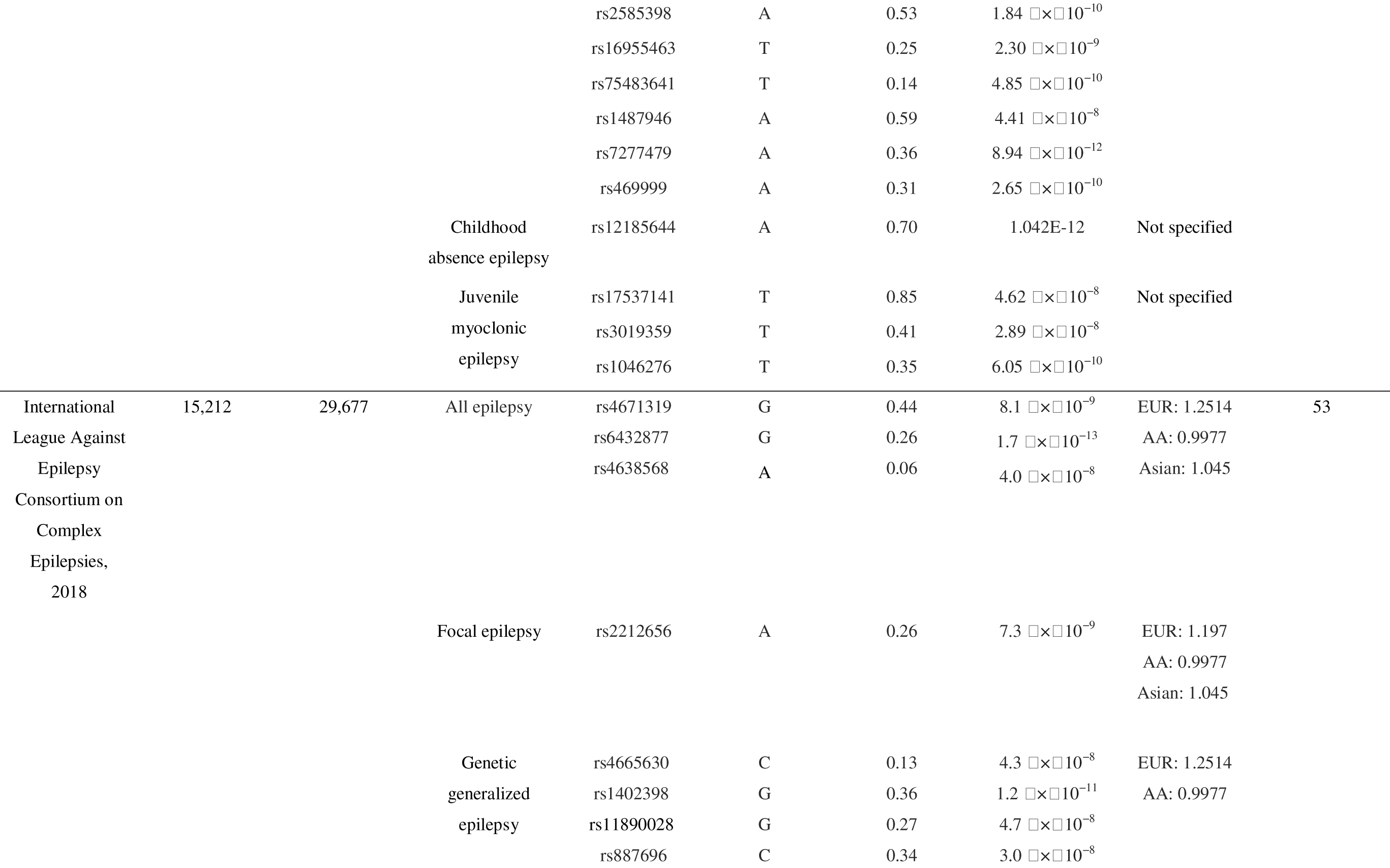
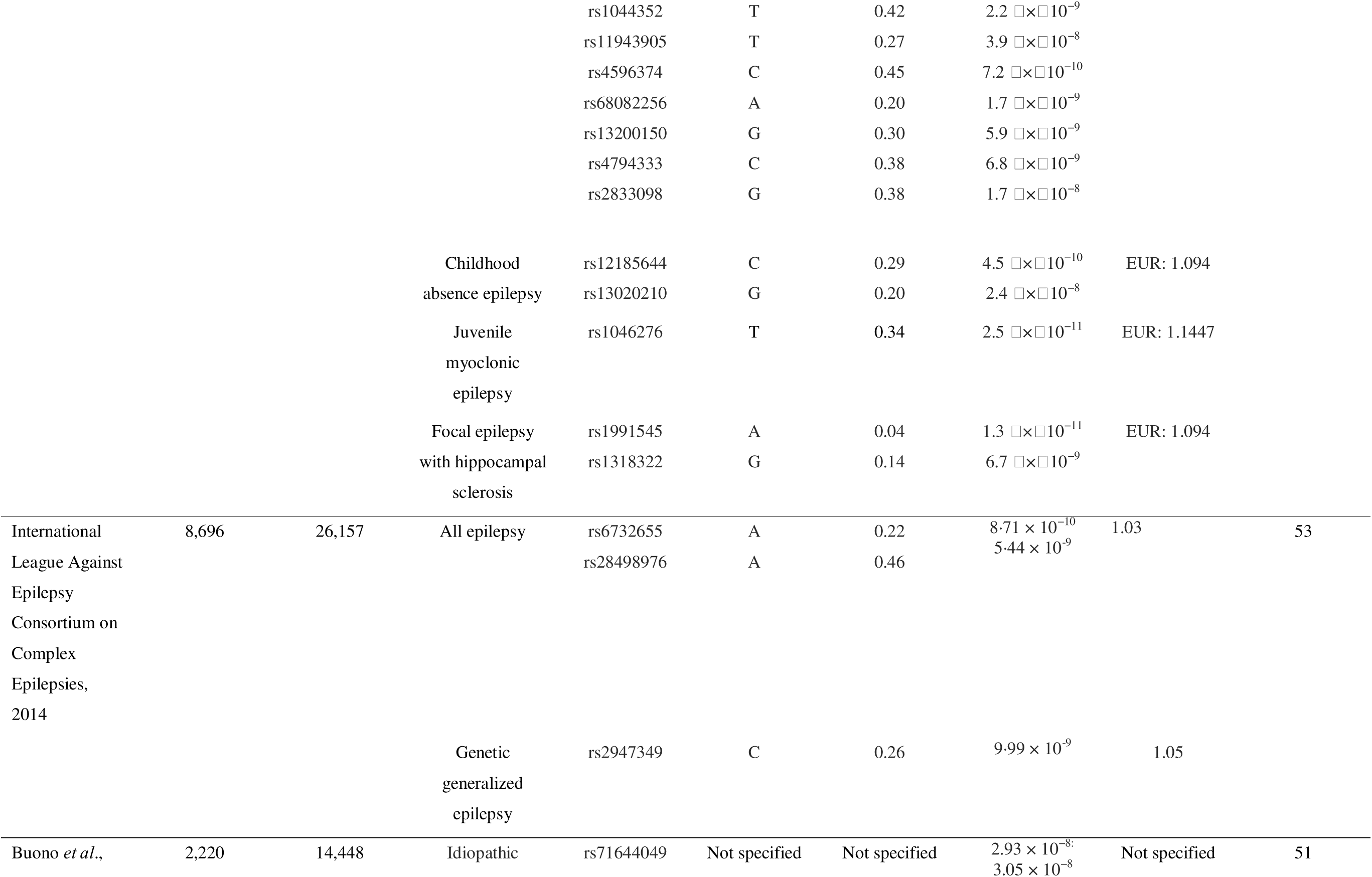
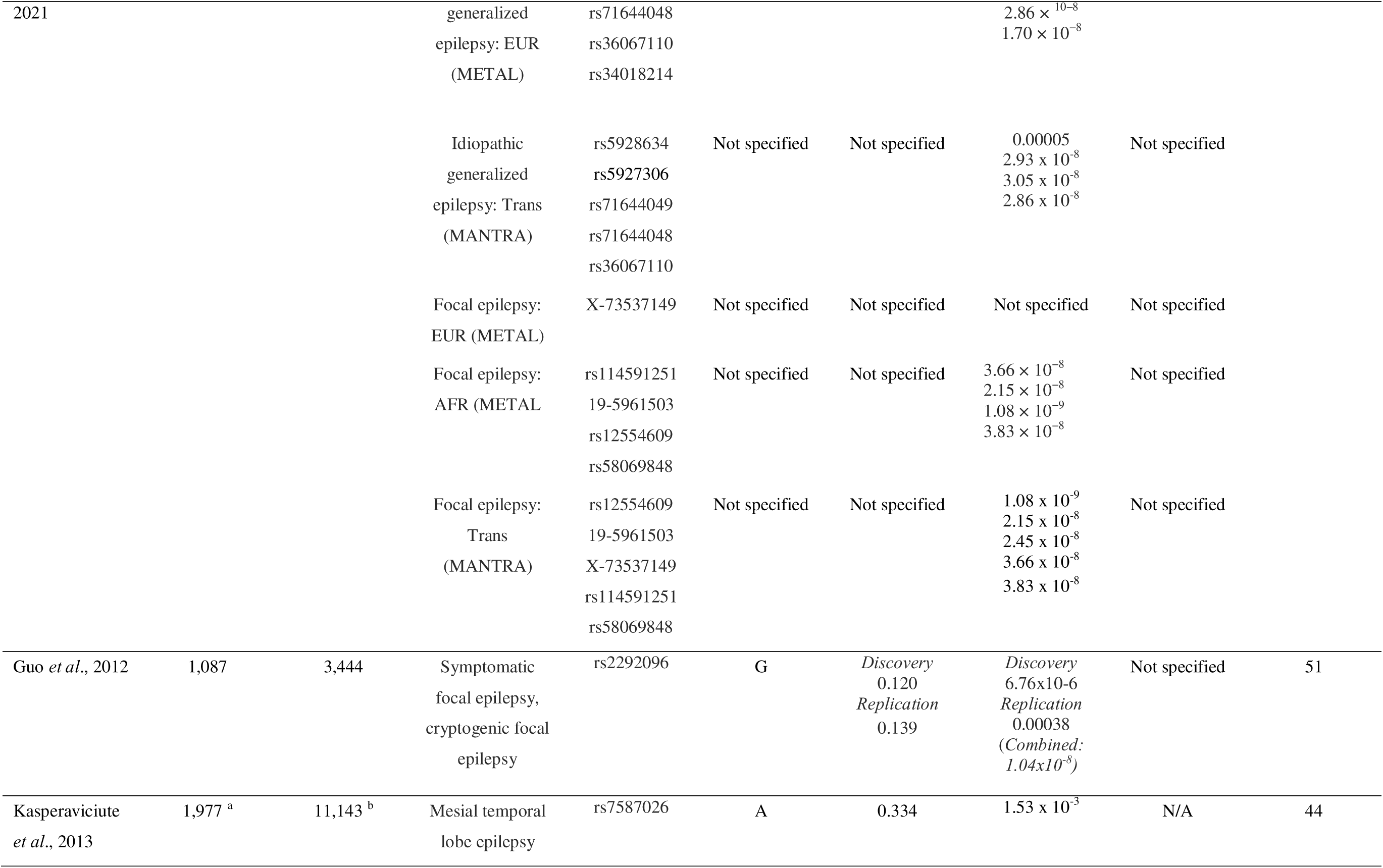
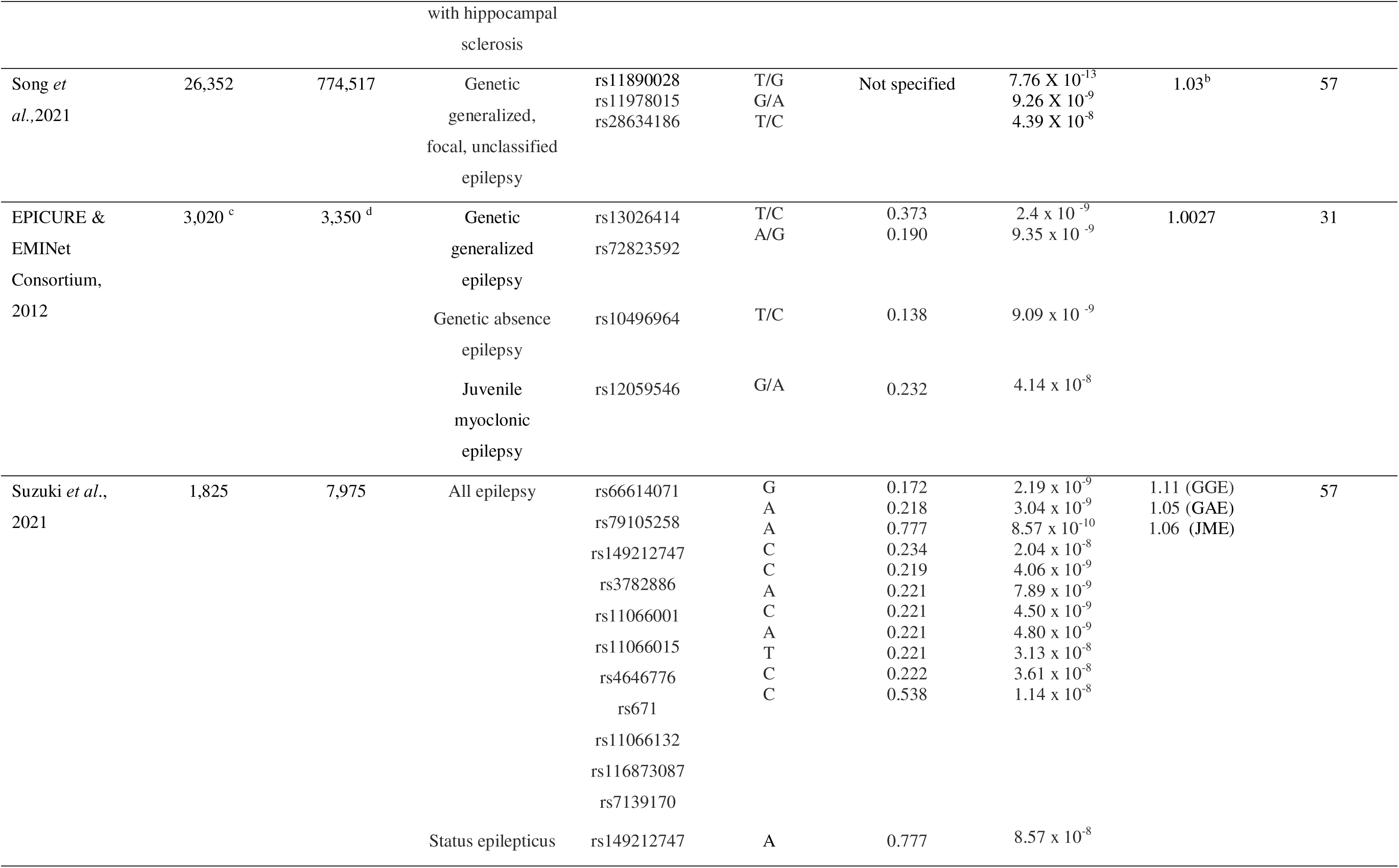
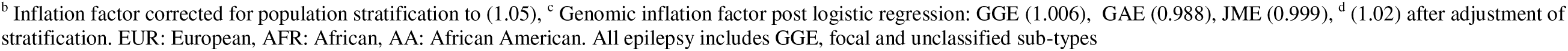
GWASs of epilepsy, with genome-wide significant SNPs only (p < 5 x 10^-8^).

The SNPs which reached genome-wide significance were grouped according to their associated epilepsy sub-type as listed in each study, with the corresponding reported candidate gene. The reported SNPs were searched on Ensembl (release 25) ^15^, with reference genome build GRCh38.14, accessed between 1-15 March 2024, to ensure consistency in reporting of SNPs and associated genes. All studies report using reference genome GRCh37. The primary function of each of the genes was established using the Uniprot database ^16^.

### 2.1 Critical appraisal of studies

To appraise the quality of the reviewed studies, the Q-Genie tool was used ^17^. This tool consists of 11 items rated on a 7-point likert scale which assess the following domains: 1) scientific basis for development of the research questions, 2) ascertainment of comparison groups (cases/controls), 3) technical and non-technical classification of genetic variants tested, 4) classification of the outcome, 5) discussion of source of bias, 6) appropriateness of sample size and power, 7) description of planned statistical analysis, 8) statistical methods applied, 9) test of assumptions used and 10) interpretation of results ^17^. The following cut-off points were used to determine the quality of each study; scores of ≤35 on the Q-Genie tool indicated poor quality, >35 and ≤45 indicated moderate quality, and >45 indicated good quality ^17^. A second independent reviewer was involved in the screening and critical appraisal as a means to decrease any possible bias, and increase accuracy of relevant studies.

## 3. Results

### 3.1 Overview of literature search

A total of 7,042 studies were identified across all databases using the search strategy described and shown in **Figure 1**. After removal of duplicates (*n* = 5,863) and entries which did not fit the inclusion criteria (*n* = 750); a final total of 429 records were uploaded to Covidence. Covidence automatically excluded an additional four duplicate records, resulting in a total of 425 records for title and abstract screening. A further 401 studies were excluded after screening. These exclusions were reviews, studies which combined epilepsy with a secondary neurological disorder or non-epileptic seizures (NES); and studies which focused on microRNA or copy number variants. From the 24 studies screened for full text, a further 13 studies were excluded due to their study design or outcome not meeting the inclusion criteria, resulting in a final total of 11 studies for full review.

The GWAS included for review were published between 2010-February 2024, with sample sizes ranging from 1,087 to 29,994 for cases and 3,444 to 774,517 for controls (**supplementary materials, Table S1.1**). The majority of studies (7 of 10) were of European ancestry, while 5 of the 10 studies included a small cohort of individuals of African ancestry. Several studies had overlapping cohorts. For example, Ishagaki *et al*., (2020) ^18^, Song *et al*., (2020) ^19^ and Suzuki *et al*., (2021) ^20^ utilized GWAS from BioBank Japan (BBJ) ^21^.

### 3.2 Quality assessment

**Table 2** shows the SNPs which reached genome wide significance (p < 5 x 10^-8^) for each study. The majority of the studies used genomic inflation as the method of quality assessment. Overall, apart from Song *et al*., (2020) ^19^ and Ishigaki *et al*., (2020) ^18^, all studies had “good global quality”, scoring >45 in Q-genie. Most studies scored between 1-2 for “other source of bias”, as they did not specifically report on such biases. Most studies scored between 1-3 for “sample size and power” as they did not include a power calculation. A detailed breakdown of each study’s scoring can be found in **supplementary material (Table S1.2).**

### 3.3 Common risk variants identified from GWAS for epilepsy

A total of 79 significant SNPs (positionally mapped to 64 genes) were identified for epilepsy across all studies (**Table 2**). From the total of 79 SNPs which reached genome-wide significance (p < 5 x 10^-8^), one SNP was identified in more than one study. Lead SNP, rs11890028, was reported in the GWAS by Song *et al*., (2021) ^19^ and ILAE (2018) ^10^, which is likely due sample overlap between these two studies. The majority of the SNPs were intronic (*n* = 41) and intergenic (*n* =23), with one synonymous (rs3782886) located in *BRAP* and one missense variant (rs671) located in *ALDH2* in Asian ancestry only. Two 3 prime UTR variants were reported, rs1047276, mapped to *ZEB2* in JME of European ancestry and rs2292096, mapped to *CAMSAP1L1* in FE of Asian ancestry. The chromosomal location of the SNPs varied, with two intergenic variants significantly associated with IGE (rs5928634 and rs5927306) located on chromosome X and mapped to the gene *TMEM47*. One SNP (rs58069848), associated with FE in both European and African ancestry groups, has not been mapped to any candidate gene.

### 3.4 Genes associated with epilepsy disorders

Several of the SNPs identified are located in ion channel genes, with sodium channel genes the most commonly reported, specifically sodium channel protein type 1 subunit alpha (*SCN1A*) which had the highest number of associated SNPs across studies (number of independent significant SNPs = 6). *SCN1A* was associated with all epilepsy types including mesial temporal lobe epilepsy with hippocampal sclerosis and febrile seizures (MTLEHS + FS). *SCN2A, SCN3A* and *SCN8A* were also found to be associated across all epilepsy types. Other reported ion channels include *GABRA2, KCNN2,GRIK1* and *SLC33A1*. A number of SNPs associated with all epilepsy types (GGE, FE and unclassified) were reported in Asian ancestry only, within the regions of 12q24.11-13, 4p15.1 and 2q21.3. Two of the SNPs (rs2292096, a 3’ UTR variant, mapped to *CAMSAP1L* and rs149212747, mapped to *LINC02355*), identified in individuals of Asian ancestry only, were associated with FE and status epilepticus, respectively.

## 4. Discussion

In this study, we systematically reviewed the results of GWASs of epilepsy. We found 11 studies with a total of 1,013,632 participants (cases = 90, 383; controls = 923, 249). The majority of these studies were comprised of participants who were predominately of European and Asian ancestry, with only a small percentage of individuals being of African ancestry. SNPs located in genes encoding ion channels, transcription factors, ribosomal protein transporters, ubiquitin ligase and protein deamination and binding are the most frequently reported to be associated with epilepsy. Two SNPs, rs2292096 and rs149212747, linked respectively to focal epilepsy (FE) and status epilepticus, were exclusively identified in individuals of Asian ancestry, alongside an Asian-exclusive synonymous variant (rs3782886) in *BRAP* and a missense variant (rs671) in *ALDH2*. We were unable to conduct a meta-analysis due to an overlap in participating cohorts in different publications ^10, 19, 21^.

Ion channel genes had the highest number of SNPs associated to epilepsy. This is in line with previous animal studies which indicated that ion channel genes account for approximately 25% of the genes identified for epilepsy ^22^, as well as with prior clinical work. These genes are grouped on the basis of the nature of their ion transport, namely voltage-gated, which includes genes *CACNA1I, CACNA2D2, KCNIP2, KCNN2, SCN1A, SCN2A, SCN3A* and *SCN8A*^23^; and ligand-gated, which includes genes *GABRA2* and *GRIK1*^24^. Ion channels play a crucial role in both the generation and propagation of neuronal action potentials in the central nervous system (CNS), therefore a disturbance in neuronal ionic flow, due to variation in an ion channel gene, may contribute towards the development of a seizure ^25^.

*SCN1A*, which is well described to be associated with epilepsy, encodes a voltage-gated sodium channel (VGSC) primarily expressed in the central nervous system (CNS) and plays a crucial role in generating and propagating action potentials in neural cells^26^. Mutations in *SCN1A* can disrupt neural activity, contributing to various forms of epilepsy, including febrile seizures (FE), unclassified epilepsy, and mesial temporal lobe epilepsy with hippocampal sclerosis and febrile seizures (MTLEHS + FS), across diverse ancestral cohorts. These findings underscore the genetic complexity underlying epilepsy, with sodium channel gene mutations having a range of different impacts on neural function and associated disorders.

*PADI6* (Peptidyl arginine deiminase 6) was mapped to several SNPs despite not having been previously associated with epilepsy. *PADI6* encodes an enzyme that deaminates proteins in molecular pathways related to epigenetic regulation of histones and autoantibody formation, suggesting that epigenetic control of gene expression may play a role in epileptogenesis ^27^. This gene was found to only be associated with IGE in individuals of European and African ancestry, with no genome-wide significant SNPs in individuals of Asian ancestry.

*FANCL* (fanconi anemia complementation group L), which encodes an ubiquitin ligase protein ^28^, also had several SNPs associated with all epilepsy and GGE. *FANCL* plays a role in the normal functioning of the immune system and CNS plasticity ^29^. In individuals with epilepsy, *FANCL* variants may lead to abnormal changes in the brain structure and function, potentially contributing to the development or maintenance of epilepsy ^31^. Two SNPs, rs3782886, rs11066001, located in the gene, *BRAP* ^28^, which also encodes a ubiquitin ligase protein, were associated with GGE, including symptomatic and unclassified epilepsy in individuals of Asian (Japanese) ancestry. Genetic variations within ubiquitin ligase protein-encoding genes, such as *FANCL* and *BRAP* may contribute to the development or persistence of epilepsy due to the impact these variations have on immune system functioning, CNS plasticity and brain structure ^31^.

SNP rs671, found to be associated with all epilepsy in Asian ancestry, is responsible for the dysfunction of aldehyde dehydrogensase type 2 (*ALDH2*), which suppresses enzymatic function ^28^. This missense variant has been associated with a number of diseases including hypertension, cancer, cardiovascular diseases and neurodegenerative diseases such as Parkinsons and Alzheimer’s ^32^. Studies focused on rs671 in recent years have focused on the association between the SNP and alcohol intake in Asian ancestry ^33^, however the role of this missense variant remains unclear. Another SNP found to be associated with all epilepsy in Asian ancestry was synonymous variant rs3782886 in breast cancer suppressor protein-associated protein (*BRAP*). Limited functional research has been done on rs3782886, with most existing studies focused on this “Asian-specific” SNP’s association to myocardial infarction and carotid atherosclerosis ^34^. Both rs671 and rs3782886 deserve further investigation to assess their association with epilepsy.

Two 3 prime UTR variants were found in European, African and Asian ancestry; namely rs1044352 and rs1046276. SNP rs1044352 was associated with GGE, mapped to protocadherin 7 (*PCDH7*) ^28^. *PCDH7* is a brain-expressed gene that shows elevated levels of expression and has been linked to CNS disorders such as epilepsy ^35^. The second SNP rs1046276 is located in calcium voltage-gated channel subunit alpha 1 *(CACNA1I*) ^28^ and has only recently been identified to be associated to JME epilepsy ^21^. Both these 3 prime UTR variants have not been well studied.

The most recent GWAS of epilepsy (ILAE, 2023) reported SNP rs62151809 to be associated to GGE in European, African and Asian ancestry. This SNP mapped to POU class 3 homeobox 3 (*POU3F3)* ^28^ which plays a role in the development of the CNS and was first reported to be associated to a neurodevelopmental disorder in 2019, specifically intellectual disability ^36^. The authors reported a missense variant of *POU3F3* in two out of thirteen individuals with mild to moderate neurodevelopmental delay, with both individuals having severe intellectual disability and epilepsy ^37^.

Several transporter channel associated genes, namely *HEATR3* (rs4638568), *TTC21B* (rs6432877, rs11890028) and *SLC33A1* (rs1991545) were found in the GWAS conducted by the International League Against Epilepsy (2018) in their trans-ethnic meta-analysis group, yet not in the GWAS conducted by Song and colleagues (2021), despite an overlap in cohorts ^9,18^. The SNPs which have been suggested to be associated to both *HEATR3* and *SLC33A1* were novel, having not been previously associated with epilepsy. The SNP rs4638568 was associated with all epilepsy groups, while rs1991545 was associated with FE only. The exact role of these genes in epilepsy disorders has not been well established and requires further investigation to better understand how variations of these genes influence the development and progression of epilepsy.

In 2022 the ILAE released updated guidelines for the classification of epilepsy syndromes. Were therefore grouped the epilepsy phenotypes according to the most recent classification by the International League Against Epilepsy in our **Table 2**. An additional challenge for reviewing data obtained from different epilepsy GWASs, is different levels of phenotypic detail are provided . Future studies would benefit from increased phenotypic harmonization.

## 5. Conclusion

Taken together, the results of this study have illustrated the value of systematically reviewing GWAS findings to prioritise risk variants for epilepsy for future follow-up investigation. Genes within ion and transport channels, in addition to genes encoding transcription factors, ubiquitin ligase and transporter proteins, in particular, should be further explored to assess their potential roles in epilepsy. Given the lack of diversity of those studied in the research reviewed here, future studies should also include participants of more diverse ancestries, particularly within African ancestries. The results of this study assist in identifying the top candidate genes and their associated SNPs for further investigation of the mechanisms underlying various epilepsy syndromes. It is notable that the majority of the SNPs and associated genes that emerged from this review are likely to be implicated in the aetiology of more than one epilepsy type, suggesting that some mechanisms are relevant across the spectrum of seizure disorders.

## Data Availability

As no primary data collection was conducted for this review, all data used in this study are publicly available and can be accessed by referring to the original sources cited in the reference list. For any additional inquiries or requests related to the data used in this systematic review, please feel free to contact the corresponding author of the publication.

## Acknowledgements

The work reported herein was made possible through funding by the South African Medical Research Council through its Division of Research Capacity Development under the SAMRC Internship Scholarship Programme. The content hereof is the sole responsibility of the authors and does not necessarily represent the official views of the SAMRC.

## Author contributions

**S. J** conceptualization, formal analysis (equal), funding acquisition, investigation, methodology, project administration, resources, validation, visualization, writing-original draft preparation; **O.W** formal analysis (equal), writing-reviewing & editing; **V I-D** supervision; writing-reviewing & editing (supporting), **L.T** supervision; writing-reviewing & editing (supporting), **D.J S**; conceptualization (supporting), supervision, writing-reviewing & editing (equal); **S.D** conceptualization (supporting), project administration (supporting), supervision (lead), validation (supporting), visualization (supporting), writing-reviewing & editing (equal)

## Declaration of conflict of interests and ethical statement

None of the authors have any conflict of interest to disclose. We confirm that we have read the Journal’s position on issues involved in ethical publication and a[rm that this report is consistent with those guidelines. This study was conducted in accordance with the Declaration of Helsinki, and the protocol was approved by the University of Cape Town Human Research Ethics Committee (183/2022).

