## supplementary material for "Systematic review of genome wide association studies (GWAS) of epilepsy identifies common risk variants and associated genes"

**Table S1.1** Detailed breakdown of all GWAS studies included in the review.

| **Study** | **Author** | **Year** | | **No. genome-wide significant SNPs** | | **Sample size** | | | **Source of sample** | **Ancestry** | | | **Genotyping platform** | **Epilepsy type** | | **Diagnosis** |
| --- | --- | --- | --- | --- | --- | --- | --- | --- | --- | --- | --- | --- | --- | --- | --- | --- |
|  |  | |  | |  | | ***Cases*** | ***Controls*** |  | |  |  | | |  |  |
| GWAS meta-analysis of over 29,000 people with epilepsy identifies 26 risk loci and subtype-specific genetic architecture | International League Against Epilepsy Consortium on Complex Epilepsies | | 2023 | | 26 | | 29,944 | 52,538 | Mixture of 24 cohorts | | Eur, Afr, As | Illumina Infinium GSA, Illumina Human610- Quadv1, Illumina 1M, Illumina Human CNV 370 duo | | | GGE, FE, UE | Clinical, EEG, MRI |
| Genome-wide mega-analysis identifies 16 loci and highlights diverse biological mechanisms in the common epilepsies | International League Against Epilepsy Consortium on Complex Epilepsies | | 2018 | | 16 | | 15,212 | 29,677 | Mixture of 24 cohorts * | | Eur, HC, AA | Affymetrix 6.0,Illumina 610 Illumina HumanCore Illumina Omni-Express-12 v1.1 Illumina Omni-Express-24 v1.1 Illumina 550 Illumina HH300 Illumina 1.2M Illumina Omni1-Quad | | | GGE, FE, UE | Clinical, EEG, MRI |
| Genetic determinants of common epilepsies: a meta-analysis of genome-wide association studies | International League Against Epilepsy Consortium on Complex Epilepsies | | 2014 | | 3 | | 8,696 | 8,696 | Mixture of 19 cohorts * | | Eur, As, Afr | Affymetrix 6.0, Illumina 610 (-quad) Illumina HumanCorE, Illumina Omni-Express-12 v1.2, Illumina Omni Express-24 v1.1, Illumina 550, Illumina HH300, Illumina 1.2M, Illumina Omni1-Quad, Illumina 660 (-quad) | | | GGE, FE, UE | Clinical, EEG, MRI |
| Genetic variation in *PADI6-PADI4* on 1p36.13 is associated with common forms of human generalized epilepsy | Buono et al. | | 2021 | | 12 | | 2,220 | 14,448 | CHOP ^e^, Nationwide Children’s Hospital, University of Pennsylvania, University of Cincinnati, University of Montreal & MGH/Harvard/Beth Israel Deaconess | | Eur, Afr | Illumina Infinium HumanHap550 Human610-Quad and HumanOmniEx-press platforms. Cohort 1, 2, 5, 6:550v1, 550v3,610 Cohort 3 and 4: OmniExpress. | | | IGE*,FE | Clinical, EEG, MRI |
| Two-stage genome-wide association study identifies variants in *CAMSAP1L1* as susceptibility loci for epilepsy in Chinese | Guo et al. | | 2012 | | 1 | | 1,087 | 3,444 | Regional hospitals in Hong Kong, University of Hong Kong & Taiwan | | HC | Illumina HumanHap 610-Quad BeadChip, Illumina HumanHap 550-Duo BeadChip | | | Symptomatic FE, cryptogenic FE | Not specified |
| Large-scale genome-wide association study in a Japanese population identifies novel susceptibility loci across different diseases | Ishigaki et al. | | 2020 | | 0 | | 2,142 | 210,310 | BJP ^f^ | | EA | Illumina HumanOmniExpress, Illumina HumanExome BeadChips | | | Not specified | Not specified |
| Epilepsy, hippocampal sclerosis and febrile seizures linked by common genetic variants around *SCN1A* | Kasperaviciute et al. | | 2013 | | 1 | | 1,977 ^a^ | 11,143 ^b^ | Austria, Belgium, USA, Finaland, Ireland, UK, Switzerland, Portugal, Netherlands, Italy | | Eur | Illumina Human610 Quadv1/Human1-2M-DuoCustom, Illumina HumanCNV370duo, llumina HumanHap300 | | | MTLE with HS | Clinical (reviewed by epileptologist) |
| Common genetic variation and susceptibility to partial epilepsies: A genome-wide association study | Kasperaviciute et al. | | 2010 | | 0 | | 3,445 | 6,935 | Mixture of 10 cohorts * | | Eur | Human610-Quadv1 (majority) HumanHap550v3, HumanHap550v1HumanHap300v1 Human1-2M-DuoCustom Human1M-Duov3, Human1Mv1 HumanCNV370-Quadv3, HumanCNV370v1 | | | Partial epilepsy (FE) | Clinical, EEG, MRI (confirmed by epileptologist) |
| Genome-wide meta-analysis identifies two novel risk for epilepsy | Song et al. | | 2021 | | 43 | | 26,352 | 774,517 | ILAE Consortium, UK Biobank Biobank Japan, FINNGEN | | Eur, As, Afr | ***ILAE*** Affymetrix 6.0, Illumina 610, Illumina HumanCore, Illumina Omni-Express-12 v1.1, Illumina Omni-Express-24 v1.2, Illumina 550, Illumina HH300 llumina 1.2M, Illumina Omni1-Quad  ***UKB*** UKB Axiom array ***BBJ*** Illumina HumanOmniExpressExome BeadChip/combination of the Illumina HumanOmniExpress, HumanExome BeadChips ***FINNGEN*** various generations of the Illumina GWAS arrays | | | GGE, FE, UE | Varied by cohort |
| Genome-wide association analysis of genetic generalized epilepsies implicates susceptibility loci at 1q43, 2p16.1,2q22.3 and 17q21.32 | EPICURE & EMINet Consortium | | 2012 | | 4 | | 3,020 ^c^ | 3,350 ^d^ | European EPICURE project | | Eur | Affymetrix Genome-Wide Human SNP Array 6.0 | | | GAE,JME | Clinical (consultant epileptologist) |
| Genome-wide association study of epilepsy in Japanese population identified an associated region at chromosome 12q24 | Suzzuki et al., | | 2021 | | 11 | | 1,825 | 7,975 | Affiliated hospitals of BBJ, Osaka-Midosuji Rotary Club, Pharma SNP Consortium | | Jap | Illumina Human OmniExpress BeadChip | | | GGE (incl. symptomatic & uncategorised) | Clinical |

^a^Total cases consisted of discovery (n=1018) and replication stage (n=959), ^b^total controls consisted of discovery (n=7552) and replication stage (n=3591). ^c^Total cases consisted of Stage 1 (n=1527), Stage 2 (n=604) and case-control group (n=889), ^d^total controls consisted of Stage 1 (n=2461) and case-control group (n=889), ^e^Jefferson, Children’s Hospital of Philadelphia , ^f^Biobank Japan Project.

*IGE includes Childhood absence epilepsy (CAE), Juvenile absence epilepsy (JAE), Juvenile myoclonic epilepsy (JME) and Generalized tonic-clonic seizures alone (GTCA) as per the updated ILAE (2017) classification.

HC-Han Chinese, AA-African American, Eur-European, EA-East Asian, As-Asian, Jap-Japanese .

GGE-Genetic generalized epilepsy, FE-Focal epilepsy, UE-Unclassified epilepsy, IGE-Idiopathic generalized epilepsy.

**Table S1.2** Q-genie risk of bias scoring for each study.

| **Study** | **Total** |
| --- | --- |
| ILAE, 2023 |  |
| ILAE, 2014 | 53 |
| ILAE, 2018 | 53 |
| Buono *et al.,* 2021 | 51 |
| Guo *et al*., 2012 | 54 |
| Ishigaki *et al*., 2020 | 44 |
| Kasperavicute *et al*., 2013 | 52 |
| Kasperavicute *et al*., 2010 | 57 |
| Song *et al*., 2021 | 31 |
| EPICURE & EMINet *et al*. 2012 | 57 |
| Suzuki *et al*., 2012 | 47 |

Scores ≤35 indicate poor quality studies, >35 and ≤45 indicate studies of moderate quality, and >45 indicate good quality studies. (All studies included control groups).
